# Machine Intelligence-Driven Forecasting for ED Triage and Dynamic Hospital Patient Routing

**DOI:** 10.64898/2026.02.18.26346566

**Authors:** Shivani Dharmavaram, Pratik Bhanushali

## Abstract

Overcrowding of emergency departments (ED) is now a problem of global health care concern due to the increase in patients. Triage systems have been established for a considerable period. However, their reliability in choosing the appropriate patient and the level of service has undergone much scrutiny. In this paper, we describe a comprehensive machine learning framework aimed at predicting critical emergency department outcomes and enabling dynamic routing decisions. Through the MIMIC-IV-ED database, which comprises more than 440,000 emergency visits, we design and assess varied predictive models, which include classical clinical scores, interpretable ML systems, classical algorithms, and deep learning architectures. We investigate three significant outcomes: hospitalization post-ED visit, critical deterioration (ICU transfer/death within 12 hours), 72-hour re-attendance in ED. The results indicate that gradient boosting algorithms can make better predictions with AUROCs of 0.820, 0.881, and 0.699 as compared to standard clinical scoring systems and complex deep learning models. The interpretable AutoScore framework which combines clinical performance with clinical transparency. We also study patterns of feature importance across prediction tasks. Moreover, we talk about how these can be implemented in realtime clinical workflows. This study builds a reproducible benchmarking platform for ED prediction research. In addition, it presents evidence-based recommendations for intelligent patient routing systems that can help enhance emergency care efficiency and resource utilization while improving patient outcomes in a high-pressure environment.

## 1 Introduction

### 1.1 Background and Motivation

Across the globe, emergency departments are the entry point to health systems. Under pressure, they deal with complex and demanding patients. The problems have worsened immensely, as emergency departments are facing unprecedented numbers and shortages in resources and staff during the COVID-19 pandemic [1]. General triage systems, and more specifically the ESI and other similar acuity-based systems, have been used to establish patient triage timeframes but have limited ability to predict exact clinical outcomes [2]. These systems greatly depend on the clinician’s ability and a few simple heuristic rules. These heuristic rules can be simple and not capture the disease trajectory due to risk factors.

The adoption of electronic health record (EHR) systems has generated numerous opportunities for data-driven clinical decision support in health care. Wide-scale, deidentified EHR databases, such as the MIMIC-IV-ED, provide rich clinical data which are temporally-structured data about vital signs, labs, medications, and outcomes [3]. Utilizing machine learning and artificial intelligence techniques can yield predictive insights from such data, which could potentially drive improved risk stratification and resource allocation in emergency settings.

### 1.2 Current Limitations and Research Gap

Despite increased interest in ML applications in emergency medicine, there are important barriers preventing the translation of these proposals into clinical practice and the comparative evaluation of different methodologies. One reason is that the absence of standardized benchmarking frameworks makes it difficult to compare different studies and algorithms directly. It is often difficult to determine whether methodological improvements significantly advance the state of the art because researchers use heterogeneous data pre-processing pipelines, variable selection criteria and evaluation metrics. In addition, much of the existing work focuses on just one prediction task or limited model architectures, providing a very incomplete perspective on relative algorithm performance across a range of relevant clinical scenarios. Finally, the trade-off between model complexity and interpretability is an important consideration for clinical implementation, but few studies assess this trade-off in the emergency department setting.

### 1.3 Research Objectives and Contributions

The shortcomings mentioned above will be addressed by this study through the following three objectives devoted to ED prediction research. First, to develop a standardized, reproducible benchmarking framework to facilitate predictive modeling research using the MIMIC-IV-ED database. Second, to comprehensively compare the performance of diverse machine learning approaches to predicting clinically relevant outcomes. Third, to investigate the impact of predictive modeling on patient routing and resource allocation in emergency care. We make available a data processing pipeline that is accessible to the public together with performance benchmarks for a range of prediction tasks. Furthermore, we will provide an analysis of feature importance that clarifies the key predictive variables. Finally, practical recommendations will be given for the clinical deployment of intelligent support systems for triage.

## 2 Related Work

The application of machine intelligence to support decision-making in complex, high-stakes environments has gained considerable traction across multiple domains, including healthcare analytics, digital forensics, and cybersecurity. Within emergency medicine, recent efforts have focused on leveraging electronic health record data to improve patient risk stratification and resource allocation [4].

A lot of research has been done on predictive modeling of emergency department outcomes using structured clinical data. Traditional triage systems like the Emergency Severity Index and various early warning scores are popular but have a limited ability to predict definite clinical trajectories. An increasing number of researchers have begun to apply machine learning approaches to identify complex, non-linear relationships between presenting features and adverse events [5].

A growing body of literature has compared conventional machine learning algorithms against deep learning architectures for clinical prediction tasks using large-scale EHR databases. Gradient boosting methods, including both extreme gradient boosting and light gradient boosting machine, have shown particular promise in tabular medical data settings. Concurrently, recurrent neural networks and representation learning techniques have been proposed to model temporal dependencies in clinical time series and diagnostic code sequences [6].

The challenge of balancing model complexity versus interpretability remains central to their clinical implementation. Although black-box models can produce better predictions, many clinicians do not trust them. Therefore, regulators are less likely to approve them. In response, several interpretable modelling frameworks have been developed that take advantage of the optimising power of machine learning while continuing to generate point-based scoring systems. The strategies target to enhance statistical performance and clinical usability [7].

Even though machine learning for the emergency department is receiving more attention, clinical translation remains hindered by important barriers. It is challenging to directly compare studies because researchers often use dissimilar data preprocessing pipelines, variable selection methods, and evaluation metrics in the absence of a standardised benchmarking framework. In addition, much of the current research focuses on a single prediction task or a limited number of models, creating an incomplete view of relative algorithm performance across clinical tasks [8].

Recent studies have also evaluated whether predictive factors can help enable dynamic routing of patients as well as resources. According to the studies, incorporating risk estimates could assist with triage, bed allocation and discharge planning within clinical workflows. There have however, been limited implementation studies and multi-centre validations in the pipeline.

Overcoming algorithm biases must ensure fairness and transparency before their implementation in the real world. Recent research highlights the requirement for comprehensive fairness audits across demographic subgroups and the design of explainability methods for emergency medicine. When considered together, these activities highlight the need for in-depth and reproducible evaluation frameworks that are clinically relevant for acute care ML-driven decision support.

## 3 Methods

### 3.1 Data Source and Study Population

This retrospective cohort was performed in the Medical Information Mart for Intensive Care IV Emergency Department (MIMIC-IV-ED) database version 2.0 imaging data. The MIMIC-IVA-ED database consists of de-identified clinical data from the emergency department of a large tertiary academic medical center in the United States [3]. The database contains details of all patients who visited the emergency department between the years 2011 to 2019 including a triage, vitals, medications, investigations, and degree of disposition. The subjects included in this study are presented in Table 1. We included all adult patients (age *≥*18 years) with complete triage documentation in our study. We excluded pediatric visits, records with missing acuity classification as well as visits that resulted in direct transfer of patient to other facilities. In total, the study enrolled 441,437 emergency department visits representing 216,877 unique patients. The data were chronologically split into training (80%, n=353,150) and test (20%, n=88,287) datasets, ensuring no overlap of patients in the two datasets to avoid leakage of information and render the assessment of model generalizability realistically.

**Table 1.**
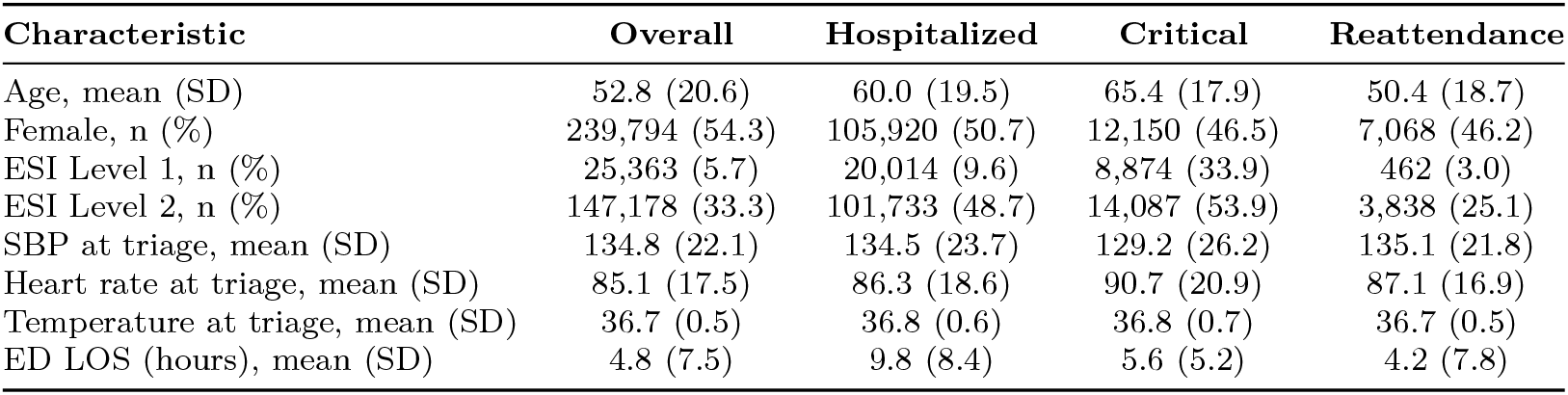
Demographic and clinical characteristics of the study cohort.

| Characteristic | Overall | Hospitalized | Critical | Reattendance |
| --- | --- | --- | --- | --- |
| Age, mean (SD) | 52.8 (20.6) | 60.0 (19.5) | 65.4 (17.9) | 50.4 (18.7) |
| Female, n (%) | 239,794 (54.3) | 105,920 (50.7) | 12,150 (46.5) | 7,068 (46.2) |
| ESI Level 1, n (%) | 25,363 (5.7) | 20,014 (9.6) | 8,874 (33.9) | 462 (3.0) |
| ESI Level 2, n (%) | 147,178 (33.3) | 101,733 (48.7) | 14,087 (53.9) | 3,838 (25.1) |
| SBP at triage, mean (SD) | 134.8 (22.1) | 134.5 (23.7) | 129.2 (26.2) | 135.1 (21.8) |
| Heart rate at triage, mean (SD) | 85.1 (17.5) | 86.3 (18.6) | 90.7 (20.9) | 87.1 (16.9) |
| Temperature at triage, mean (SD) | 36.7 (0.5) | 36.8 (0.6) | 36.8 (0.7) | 36.7 (0.5) |
| ED LOS (hours), mean (SD) | 4.8 (7.5) | 9.8 (8.4) | 5.6 (5.2) | 4.2 (7.8) |

The overall methodological framework of this paper which includes the preprocessing of data, the formation of the prediction task, model development, evaluation, and clinical implementation is illustrated in Figure 1.

**Fig. 1.**
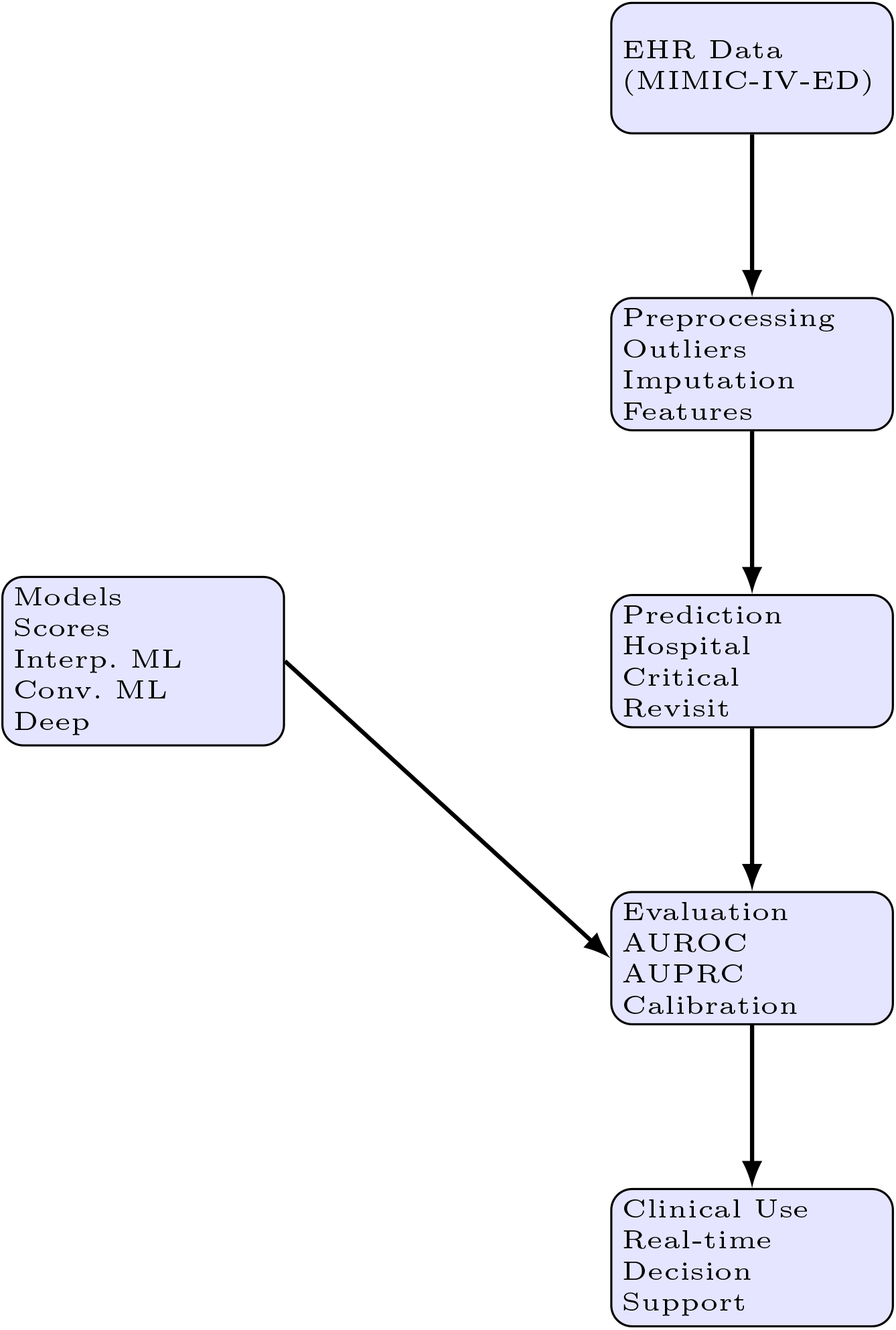
Overview of the predictive modeling framework.

### 3.2 Prediction Tasks and Outcome Definitions

We created three clinically relevant prediction tasks using consensus guidelines and the literature.

#### Hospitalization Prediction

Hospitalization was defined as admission to inpatient care (including observation status) following emergency department evaluation. Patients who left to drive home, transfer to another facility, or left without discharge were considered non-hospitalized. This result indicates the severity of patients and how the services are used.

#### Critical Outcome Prediction

Critical Outcome Prediction was defined as a composite endpoint consisting of either inpatient death or transfer to intensive care or cardiac care a case of a 12-hour period after arrival.

#### 72-Hour Reattendance Prediction

This endpoint defines the patients who are deteriorating quickly and who need action and resource mobilization.

We defined reattendance as a non-scheduled visit to the emergency department for the same medical issue within 72 hours of discharge from the index visit. This measure is a suggestion of a possible lack of quality in Initial assessment, treatment or discharge planning.

### 3.3 Feature Engineering and Data Preprocessing

Clinical variables from multiple relational tables in MIMIC-IV-ED were extracted and summarized at the visit level. These features were divided into five categories: (1) demographic characteristics (age, sex); (2) triage information (vital signs, chief complaint, ESI level); (3) historical utilization (prior ED visits, hospitalization, ICU admissions); (4) comorbidity indices (Charlson and Elixhauser scores that were made from the ICD codes); and (5) ED course features (vital signs trends, medication given, length of stay).

A multi-stage imputation technique was adopted to handle missing data. We treated physiological outliers (e.g. SpO_2_ > 100%, heart rate < 30 bpm) as missing values using clinically plausible ranges. We imputed continuous variables as the median of the training set and categorical variables as the mode. Common time intervals were used to align the temporal variables (for example, serial vital measurements) and statistical aggregates (mean, minimum, maximum, trend slope) were summarised.

### 3.4 Predictive Modeling Approaches

We executed and contrasted four classifications of prediction models.

#### Traditional Clinical Scoring Systems

Established clinical scores were calculated directly from input variables without model training: Emergency Severity Index (ESI), Modified Early Warning Score (MEWS), National Early Warning Score (NEWS and NEWS2), Rapid Emergency Medicine Score (REMS), and Cardiac Arrest Risk Triage (CART).

#### Interpretable Machine Learning

The AutoScore framework was designed to generate integer-based clinical scores through automated variable selection, weighting, and cut-point optimization in [9]. This method ensures full interpretability and leverages machine learning for optimal feature engineering.

#### Conventional Machine Learning Algorithms

Three commonly used supervised algorithms were implemented: logistic regression with L2 regularization, random forest with 100 decision trees, and gradient boosting with 100 sequential estimators. Unless otherwise specified, all models were implemented using *scikit-learn* with default hyperparameters. Before training, input features were standardized for numerical stability and fair comparison between models [10]. Cross-validation was employed to evaluate model performance with stratification in order to reduce overfitting. The linear baseline is logistic regression. Similarly, the ensemble-based random forest and gradient boosting models capture interaction between features and complex decision boundaries. The performances of these established techniques serve as a significant yardstick and the benchmark for our proposals.

#### Deep Learning Architectures

An experiment was conducted on three architectures: Multilayer perceptron (MLP) with two hidden layers, LSTM networks for temporal data, and Med2Vec for representation learning of diagnostic codes [11]. The models were executed with Keras utilizing Adam optimization and early stopping [12].

### 3.5 Model Training and Evaluation

All models were trained exclusively on the training subset using five-fold crossvalidation for hyperparameter tuning. Model performance was evaluated on the held-out test subset using area under the receiver operating characteristic curve (AUROC) as the primary metric. Additional metrics included area under the precision-recall curve (AUPRC), sensitivity, specificity, positive predictive value, and negative predictive value. Confidence intervals (95%) were calculated using 1000 bootstrap samples. Computational efficiency was assessed through training time measurement on standardized hardware.

## 4 Results

### 4.1 Model Performance Across Prediction Tasks

All the performance metrics for all the models on all tasks are as shown in Table 2. The performance of gradient boosting was consistently strong with gradients of AUROC (95% CI) equal to 0.820 (0.818-0.823), 0.881 (0.877-0.886) and 0.699 (0.689-0.712) for hospitalization, critical outcome, and reattendance respectively. The Clinical Use Real-time Decision Support multilayer perceptron (MLP) architecture showed superior performance over gradient boosting for important outcome prediction (AUROC 0.883, 95% CI: 0.880-0.888) without statistical significance.

**Table 2.** Comparative performance of predictive models across three clinical outcomes.

| Model | AUROC (95% CI) | AUPRC (95% CI) | Sensitivity | Specificity | Time(s) | Variables |
| --- | --- | --- | --- | --- | --- | --- |
| **Hospitalization Prediction** |  |  |  |  |  |  |
| Gradient Boosting | 0.820 (0.818-0.823) | 0.794 (0.791-0.798) | 0.743 | 0.743 | 62 | 64 |
| MLP | 0.823 (0.822-0.826) | 0.797 (0.794-0.800) | 0.759 | 0.735 | 62 | 64 |
| Random Forest | 0.819 (0.818-0.822) | 0.786 (0.784-0.789) | 0.754 | 0.734 | 58 | 64 |
| AutoScore | 0.793 (0.791-0.797) | 0.756 (0.753-0.760) | 0.722 | 0.721 | 170 | 10 |
| ESI | 0.711 (0.709-0.714) | 0.632 (0.628-0.636) | 0.582 | 0.784 | 0 | 1 |
| NEWS | 0.581 (0.579-0.584) | 0.555 (0.552-0.559) | 0.565 | 0.540 | 0 | 6 |
| **Critical Outcome Prediction** |  |  |  |  |  |  |
| MLP | 0.883 (0.880-0.888) | 0.386 (0.375-0.404) | 0.810 | 0.796 | 376 | 64 |
| Gradient Boosting | 0.881 (0.877-0.886) | 0.388 (0.374-0.405) | 0.801 | 0.799 | 76 | 64 |
| AutoScore | 0.846 (0.842-0.851) | 0.278 (0.267-0.293) | 0.804 | 0.728 | 166 | 7 |
| ESI | 0.804 (0.801-0.809) | 0.194 (0.187-0.205) | 0.870 | 0.640 | 0 | 1 |
| CART | 0.707 (0.701-0.713) | 0.141 (0.132-0.148) | 0.590 | 0.731 | 0 | 4 |
| NEWS | 0.634 (0.627-0.640) | 0.141 (0.132-0.144) | 0.464 | 0.795 | 0 | 6 |
| **72-Hour Reattendance Prediction** |  |  |  |  |  |  |
| Gradient Boosting | 0.699 (0.689-0.712) | 0.162 (0.149-0.177) | 0.653 | 0.631 | 30 | 67 |
| MLP | 0.696 (0.687-0.709) | 0.160 (0.146-0.174) | 0.625 | 0.652 | 93 | 67 |
| LSTM | 0.697 (0.683-0.712) | 0.158 (0.144-0.171) | 0.637 | 0.657 | 10,454 | 67 |
| AutoScore | 0.673 (0.665-0.684) | 0.114 (0.107-0.124) | 0.621 | 0.628 | 180 | 12 |
| Logistic Regression | 0.683 (0.677-0.697) | 0.155 (0.141-0.169) | 0.620 | 0.642 | 3 | 67 |

There was a substantial loss in the discriminative ability of traditional clinical scoring systems. The Emergency Severity Index (ESI) had AUROCs of 0.711 for admission to hospital and 0.804 for critical, with other scores (MEWS, NEWS, REMS, CART) performing even worse. The interpretable AutoScore framework produced competitive performance with AUROCs of 0.793, 0.846, and 0.673 – only modest reductions relative to best-performing black-box models.

Interestingly, more sophisticated deep learning architectures (LSTM, Med2Vec) proved much less effective than simpler machine learning ones despite their greater sophistication and compute requirements. This means that simple algorithms may capture the predictive signal in the structured emergency department data without complex neural network designs.

Comparative performance of predictive models across three clinical outcomes desctibed in table 2

### 4.2 Feature Importance Analysis

As demonstrated in Figure 2, the clinical variables exhibited varying levels of importance in the context of different prediction tasks. This was determined through the application of random forest feature importance. According to the prediction of hospitalization variable, age was the most important (importance 0.123) followed by triage acuity (ESI level 0.112) and systolic blood pressure at triage (0.086). In predicting critical outcomes, age had the highest ranking (0.101) followed by systolic blood pressure (0.095) and heart rate (0.094). The predictive landscape in the reattendance case was seen to be very different with emergency department length of stay being the most important feature (0.084), and age (0.084) and systolic blood pressure during ED stay (0.079).

**Fig. 2.**
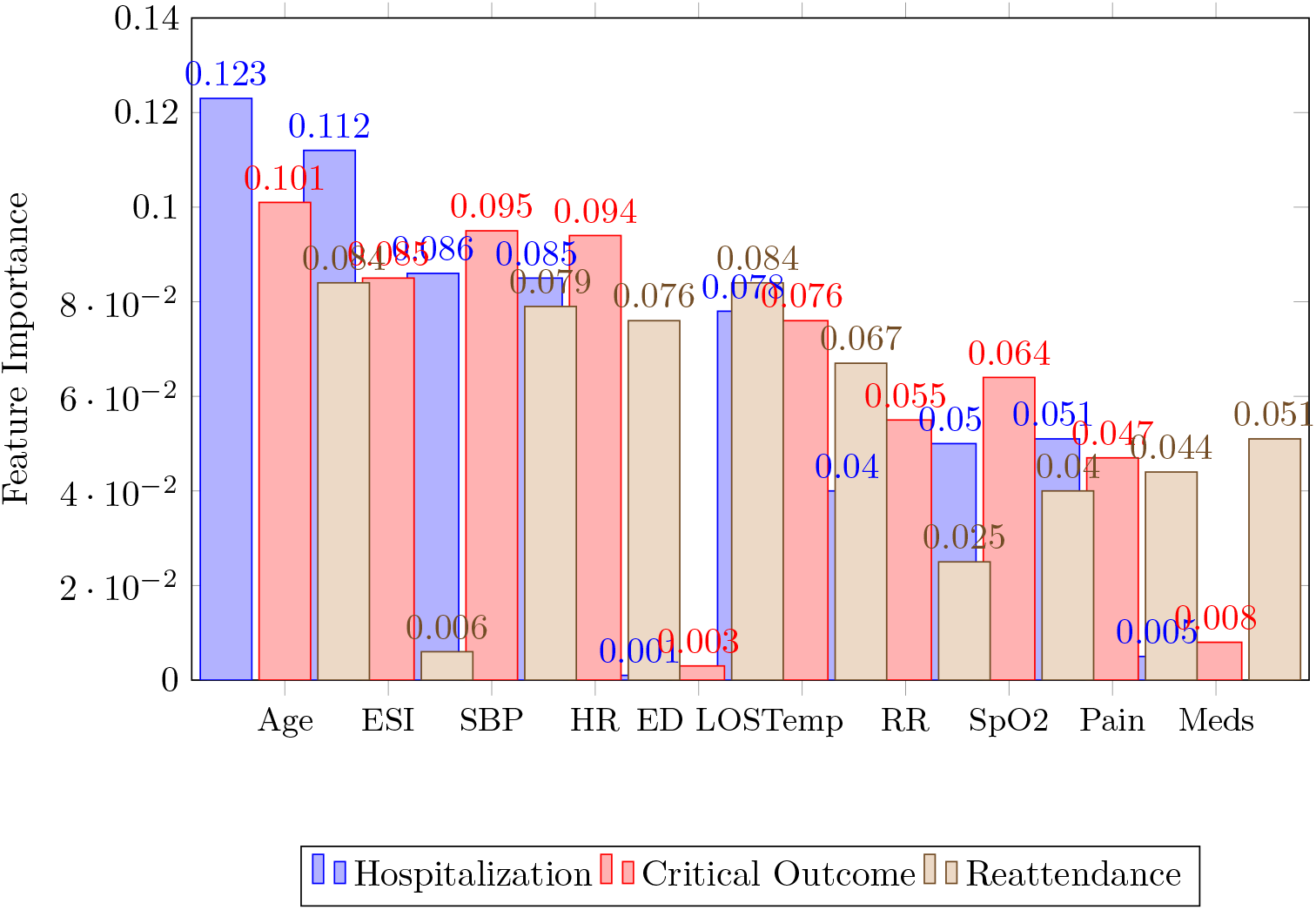
Feature importance rankings across prediction tasks (top 10 variables shown)

The results indicate different clinical signatures for different prediction tasks. Though both hospitalization and critical outcome prediction primarily use traditional acuity indicators (age, vital signs, triage level), reattendance prediction seems primarily reliant on process indicators (ED LOS, medication administration pattern) and dynamic change in physiology in ED course.

### 4.3 Calibration and Clinical Utility Analysis

We performed reliability curves to measure calibration and quantified the decision curve analysis across probability thresholds with clinical utility beyond discrimination metrics. The hospitalization prediction utilizing the gradient boosting and MLP model showed excellent calibration with the predicted probabilities closely aligning to the observed event rates in the risk deciles. All models demonstrated some level of overestimation in low-risk groups and underestimation in high-risk groups, albeit gradient boosting displayed the best calibration overall for critical outcome prediction.

Critical outcomes for various types were performed for the selected models by receiver operating characteristic curves which have a graphical representation in Fig. 3

**Fig. 3.**
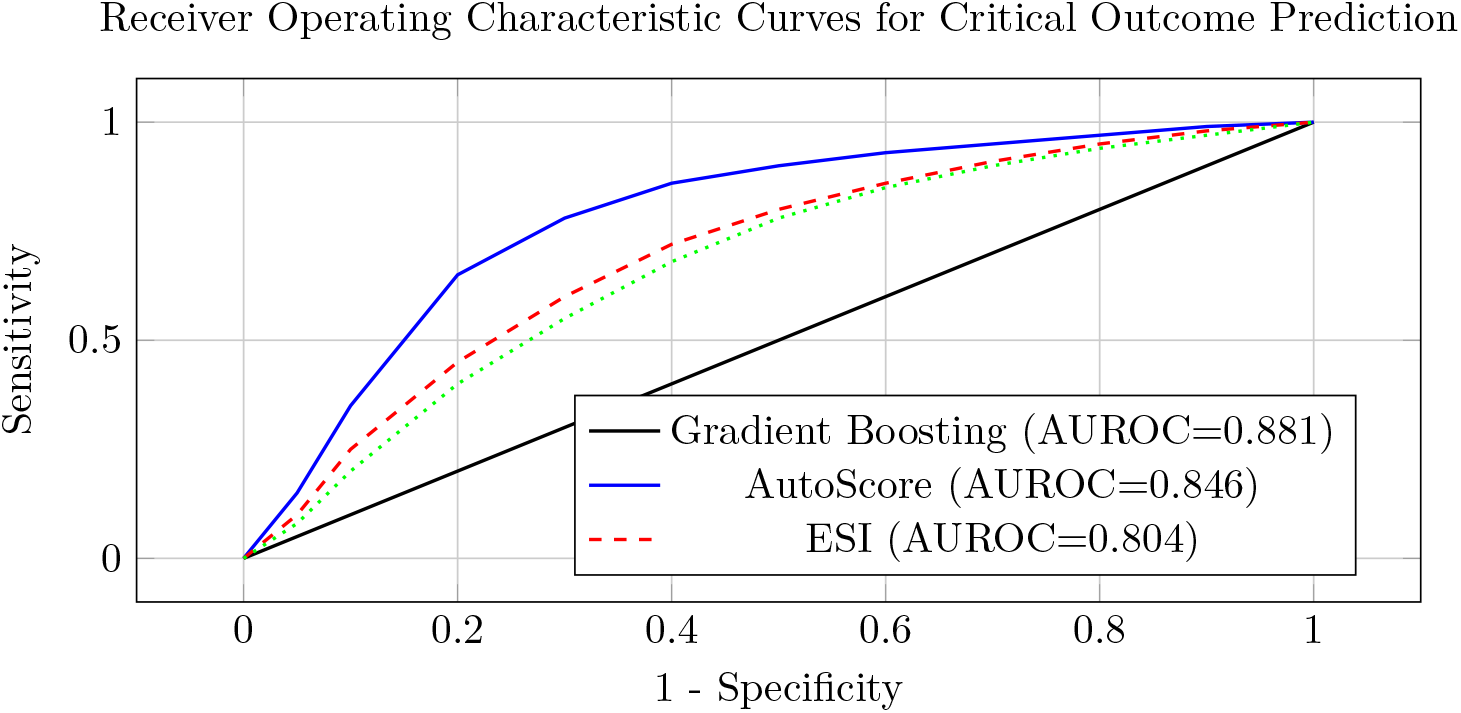
ROC curves comparing top-performing models for critical outcome prediction.

The machine learning models assessed via decision curve analysis provided a positive net benefit across clinically important probability thresholds for hospitalization and critical outcome. For reattendance prediction, its clinical usefulness was more restricted owing to lower overall discriminative ability with positive net benefit restricted to narrow threshold ranges.

## 5 Discussion

### 5.1 Interpretation of Key Findings

We identified several key findings from our comprehensive benchmarking study for prediction research and clinical implementation in the ED. Initially, all predictive tasks demonstrated that there is a consistently superior performance of machine learning algorithms compared to standard clinical scoring systems. It builds on the evidence of earlier dedicated studies and also validates these earlier studies with rigorous comparisons of multiple models. The performance gain is particularly notable in the prediction of critical outcomes, with gradient boosting and MLP models attaining AUROCs above 0.880, compared with 0.804 for ESI and 0.634 for NEWS.

The limited performance gain of complex deep learning architectures over standard ML algorithms suggests that structured emergency department data may not have sufficiently complex signal properties that justify elaborate neural network designs. This finding is in line with recent literature that questions the necessity of deep learning in the predictive tasks on tabular medical data [13]. The heavy computation requirements of the LSTM and Med2Vec models also limit their practical appeal given marginal performance gains.

The trade-off between accuracy and interpretability is secondly important for clinical implementation. Although black-box models (_gradient boosting, MLP_) produce the highest discriminative performance, their lack of interpretability prevents clinicians from trusting them and hinders regulatory approval. The AutoScore framework strikes a compelling balance by delivering interpretable integer scores with only small performance losses. This may be the right balance for real-world clinical applications, particularly in emergency medicine which requires rapid decision-making and rationale.

### 5.2 Clinical Implications for Patient Routing

The predictive models created in the study may be useful in the patient routing and resource allocation in emergency care. Three defined routes of implementing warranted.

#### Real-Time Triage Decision Support

Real-time decision assistance for allocation. Hospitalisation and critical outcome predictions may be integrated into electronic triage interfaces to facilitate the provision of more objective risk estimates to clinicians than clinical judgment alone. Upon a prediction of high-risk, it is likely that a doctor’s assessment and diagnostic testing will happen sooner, and a bed will be reserved early. Systems must aid clinical expertise and not substitute it. The level of uncertainty associated with the prediction should be clearly displayed, along with evidence indicating which features are contributing to its prediction.

#### Dynamic Resource Allocation

Dynamic Resource Care needs predictions can be used to inform real-time decisions about staffing and resource allocation in the care environment. Emergency departments are encouraged to have a tiered response protocol. To enable the preventive activation of a team of critical care nurses and respiratory therapists, with the preparation of resuscitation equipment, prior to the clinical deterioration of patients. The prediction of hospital admissions may be useful to the inpatient unit in bed allocation.

#### Discharge Planning Optimisation

For discharge planning optimisation, those likely to bounce back at discharge can be identified and intervened to reduce post-discharge reattendance. High-risk patients might receive additional info on discharge, a follow-up appointment and transitional care. Merging with pharmacy systems might help manage drug reconciliation and support compliance for patients with complex medications.

### 5.3 Limitations and Future Directions

This study has a number of limitations. The MIMIC-IV-ED database is a singlecenter database, which may limit its generalizability to other health systems that differ in patient populations, clinical practices, and documentation standards. Validation by other health systems is an important next step before use in the clinic. In addition, our analysis focuses solely on structured EHR data rather than using the wealth of information found in clinical notes, imaging studies, and real time streams of physiological monitoring data. Combining various data sources through multimodal learning approaches might result in extra predictive gains.

The retrospective observational design prevents the evaluation of causality and effects of treatment. The implementation studies of predictive model need to be carried out in the future to emphasise clinical outcomes, workflow as well as resource impact. The fourth point highlights issues related to fairness, transparency, and bias algorithms that need to be addressed before actual clinical deployment. Future studies need to perform thorough fairness reviews on breakdowns along demographic lines, in addition to developing explainability anti-money tools that are fit for purpose in the context of emergency medicine The receiver operating characteristic curves are shown in Figure 3 for selected models and used to predict critical outcome.

### 5.4 Practical Implementation Considerations

For an emergency department’s predictive model to be clinically useful, there are many practical considerations. It must integrate seamlessly using current EHR systems with little disruption to clinical workflows. Prediction display interfaces need to show concise information but should also allow access to supporting information. Having clinician education and engagement throughout implementation is important in building trust and use. As clinical practices continue to evolve, there will be a need for continuous monitoring and periodic model retraining to ensure proper performance. Compliance with regulations is very important. The use of predictive models to support clinical decision-making may require regulatory approval, depending on the jurisdiction and intended use case of the model itself. The public reporting of all aspects of model development, validation, and limitations will be essential for regulatory applications and clinician understanding. Prior to implementation, accountability mechanisms and appeal processes should be established for algorithm governance.

## 6 Conclusion

This research presents a complete benchmarking framework for machine learning prediction in the emergency department (ED), and demonstrates that the gradient boosting and multilayer perceptron models outperform traditional clinical scoring systems on three major prediction tasks. The interpretable AutoScore framework strikes a pragmatic balance between predictive performance and clinical interpretability, representing a promising avenue for real-world implementation. Analyzing feature importance uncovers unique clinical signatures for diverse prediction tasks, guiding intervention development.

Bringing intelligent prediction systems into workflows of the emergency department (ED) can improve patient routing, resource allocation and clinical outcomes. Future studies should concentrate on prospective implementation studies, multimodal data integration, multi-centre validation and ethical framework development. Due to increased usage and resource restrictions in emergency departments, it would be seen that use of analytics for decision support systems are becoming an increasingly important aspect for providing quality emergency care in an efficient manner.

## Data Availability

The data used in this study are derived from the publicly available MIMIC-IV-ED database, accessible via the PhysioNet repository after completing required credentialing and data use agreements. No new data were generated for this study.

https://physionet.org/content/mimic-iv-ed/

## References

[1] Jeffery, M.M., D’onofrio, G., Paek, H., Platts-Mills, T.F., Soares III, W.E., Hoppe, J.A., Genes, N., Nath, B., Melnick, E.R.: Trends in emergency department visits and hospital admissions in health care systems in 5 states in the first months of the covid-19 pandemic in the us. JAMA Internal Medicine 180(10), 1328–1333 (2020)

[2] Eitel, D.R., Travers, D.A., Rosenau, A.M., Gilboy, N., Wuerz, R.C.: The emergency severity index triage algorithm version 2 is reliable and valid. Academic Emergency Medicine 10(10), 1070–1080 (2003)

[3] Johnson, A., Bulgarelli, L., Pollard, T., Celi, L., Mark, R., Horng, S.: MIMIC-IV-ED. PhysioNet (2021)

[4] El Arab, R.A., Al Moosa, O.A.: The role of ai in emergency department triage: an integrative systematic review. Intensive and Critical Care Nursing 89, 104058 (2025)

[5] Sax, D.R., Warton, E.M., Mark, D.G., Reed, M.E.: Emergency department triage accuracy and delays in care for high-risk conditions. JAMA Network Open 8(5), 258498 (2025)

[6] Joseph, J.W., Kennedy, M., Landry, A.M., Marsh, R.H., Baymon, D.M.E., Im, D.E., Sánchez, L.D.: Race and ethnicity and primary language in emergency department triage. JAMA Network Open 6(10), 2337557 (2023)

[7] Janerka, C., Leslie, G.D., Gill, F.J.: Patient experience of emergency department triage: an integrative review. International Emergency Nursing 74, 101456 (2024)

[8] Stewart, J., Lu, J., Goudie, A., Arendts, G., Meka, S.A., Freeman, S., Dwivedi, G.: Applications of natural language processing at emergency department triage: A narrative review. PLoS One 18(12), 0279953 (2023)

[9] Xie, F., Chakraborty, B., Ong, M., Goldstein, B., Liu, N.: AutoScore: A machine learning-based automatic clinical score generator and its application to mortality prediction using electronic health records. JMIR Medical Informatics 8(10), 21798 (2020)

[10] Pedregosa, F., Varoquaux, G., Gramfort, A., Michel, V., Thirion, B., Grisel, O., Blondel, M., Prettenhofer, P., Weiss, R., Dubourg, V., et al.: Scikit-learn: Machine learning in Python. Journal of Machine Learning Research 12, 2825–2830 (2011)

[11] Choi, E., Bahadori, M.T., Searles, E., Coffey, C., Thompson, M., Bost, J., Tejedor-Sojo, J., Sun, J.: Multi-layer representation learning for medical concepts. In: Proceedings of the 22nd ACM SIGKDD International Conference on Knowledge Discovery and Data Mining, pp. 1495–1504 (2016)

[12] Gulli, A., Pal, S.: Deep Learning with Keras. Packt Publishing Ltd, ??? (2017)

[13] Xie, F., Yuan, H., Ning, Y., Saffari, S.E., Chakraborty, B., Liu, N.: Deep learning for temporal data representation in electronic health records: A systematic review of challenges and methodologies. Journal of Biomedical Informatics 126, 103980 (2022)

